# Community incidence patterns drive the risk of SARS-CoV-2 outbreaks and alter intervention impacts in a high-risk institutional setting

**DOI:** 10.1101/2022.11.22.22282480

**Authors:** Sean M. Moore, Guido España, T. Alex Perkins, Robert M. Guido, Joaquin B. Jucaban, Tara L. Hall, Mark E. Huhtanen, Sheila A. Peel, Kayvon Modjarrad, Shilpa Hakre, Paul T. Scott

**Affiliations:** Department of Biological Sciences and Eck Institute for Global Health, University of Notre Dame, Notre Dame, Indiana 46556; Moncrief Army Health Clinic, Fort Jackson, South Carolina 29207; United States Army Training Center, Fort Jackson, South Carolina 29207; Diagnostics and Countermeasures Branch, Walter Reed Army Institute of Research, Silver Spring, Maryland 20910; Emerging Infectious Diseases Branch, Walter Reed Army Institute of Research, Silver Spring, Maryland 20910; Henry M. Jackson Foundation for the Advancement of Military Medicine, Inc., Bethesda, Maryland 20817

**Keywords:** SARS-CoV-2, COVID-19, agent-based model, epidemiology, disease dynamics, high-risk settings, forecasting

## Abstract

Optimization of control measures for severe acute respiratory syndrome coronavirus 2 (SARS-CoV-2) in high-risk institutional settings (e.g., prisons, nursing homes, or military bases) depends on how transmission dynamics in the broader community influence outbreak risk locally. We calibrated an individual-based transmission model of a military training camp to the number of RT-PCR positive trainees throughout 2020 and 2021. The predicted number of infected new arrivals closely followed adjusted national incidence and increased early outbreak risk after accounting for vaccination coverage, masking compliance, and virus variants. Outbreak size was strongly correlated with the predicted number of off-base infections among staff during training camp. In addition, off-base infections reduced the impact of arrival screening and masking, while the number of infectious trainees upon arrival reduced the impact of vaccination and staff testing. Our results highlight the importance of outside incidence patterns for modulating risk and the optimal mixture of control measures in institutional settings.

**Disclaimer:** The views expressed are those of the authors and should not be construed to represent the positions of the U.S. Army, the Department of Defense, or the Henry M. Jackson Foundation for the Advancement of Military Medicine, Inc.

## Introduction

Over the course of the SARS-CoV-2 pandemic there have been numerous outbreaks linked to indoor environments where close-contact interactions make preventing transmission difficult (*1*–*3*). SARS-CoV-2 is highly transmissible in high-density, indoor settings where prolonged exposures are common, such as on cruise ships and naval vessels as well as in overnight camps, prisons, and nursing homes (*1, 2, 4*–*6*). Basic training camps for new military recruits are particularly high-risk settings due to the frequent arrival of new individuals and the close contacts between trainees including group sleeping quarters (*7, 8*). The average number of contacts and the probability of transmission per contact may both be higher in military basic training and similar institutional settings than they are in the broader community, leading to a higher basic reproduction number (R_0_) (*9*). In addition to R_0_, another critical factor determining the risk of an outbreak in these settings is the probability or rate of SARS-CoV-2 introductions from the outside community (*10*–*12*). However, the relative importance of external importation pressure versus local transmission dynamics for outbreak risk in high-transmission institutional settings is not well understood. The relative importance of these two factors has important implications for outbreak prevention and control, because interventions can vary in their effectiveness at preventing introductions versus controlling spread of transmission once SARS-CoV-2 has already been introduced into a setting.

The importation risk into high-risk institutional settings has likely varied across both time and location over the course of the pandemic. Temporal patterns of COVID-19 incidence at the local- and national-levels have also varied over the course of the pandemic, with several large national waves of incidence (*13*). In addition, the timing and magnitude of peak incidence at the state or county level has fluctuated over the course of these waves. There have also been regional outbreaks outside of these waves (*14*). To assess the risk of an outbreak in high-risk settings, it is therefore important to estimate the importation risk based on the current epidemiological context of the broader community in which the setting is situated. At one extreme, without any introductions the outbreak risk is zero, regardless of R_0_; while, if multiple infectious individuals are introduced into a large susceptible population with a sufficiently high R_0,_ an outbreak is highly likely (*15*). If, however, R_0_ is closer to or below one, then local conditions and interventions within the institutional setting are likely an important component of the outbreak risk in that setting.

If outbreak risk is influenced by external importation pressures, then assessing the risk of introductions in real time, or in advance, is a useful component of outbreak prevention and control in institutional settings. As a result, it is important to determine the extent to which the expected number of introductions or importations into a particular setting can be predicted from reported incidence at the local or regional level. Examining the association between national incidence and viral introductions into military training camps is essential, as new recruits arrive on a weekly basis from all over the United States. Beyond the immediate importation pressure, the risk of an outbreak will also depend on the infection history and vaccination status of the population within the setting, as these variables determine an individual’s susceptibility to infection (*13*). In the absence of serology testing, it is possible that population-level susceptibility in a particular setting at any given time could be estimated from local- or national-level infection and vaccination trends, assuming that these are representative of the population of interest (*16*). It is therefore an urgent question whether publicly available incidence and vaccination data, as well as estimates of important geographic or demographic variations in this data, could be used to estimate the probability of introduction into a particular setting, as well as the likely attack rate or outbreak size following an introduction(s).

The effectiveness of control measures in high-risk institutional settings will also depend on both the importation risk and R_0_ (*4, 17, 18*). If introduction patterns are the primary driver of outbreak risk then arrival screening and other interventions that prevent importations are important. However, if local conditions, particularly the setting-specific R_0_ or transmission rates, are the main driver of outbreak risk, then interventions that prevent local transmission within the community are critical. Active surveillance testing is likely to be important in settings where new individuals regularly arrive from the broader community and serve as potential vectors of SARS-CoV-2 into the local community (*12, 19*–*21*). Regarding local conditions, congregate living settings, such as prisons, overnight summer camps, naval vessels, and military training bases, make it difficult to implement certain social distancing measures and therefore may require other interventions (*22*–*26*). Non-pharmaceutical interventions (NPIs), such as mask wearing or testing-and-isolation strategies, that lower R_0_ below one on their own in low- or moderate-risk settings may not prevent an outbreak in a setting with a high intrinsic R_0_ value (*26*). As a result, a mix of interventions should be considered in these settings. Given the limitations of different control measures, it is important to understand which combination of interventions is most likely to be effective in a particular setting, and whether their effectiveness changes over time with shifts in SARS-CoV-2 epidemiology.

In this study, we extended an earlier analysis of outbreak dynamics in a military training camp to assess the relative importance of the impact of transmission dynamics in the broader community on importation pressure and outbreak risk during training. First, we tested the predictability of importation pressure in this setting by using publicly available infection estimates to predict SARS-CoV-2 introductions into camps from newly arrived military recruits from all over the United States, as well the probability that military trainers and support staff were infected off base over the course of the training camp. Next, we used an individual-based transmission model to estimate R_0_ in this high-risk setting while taking into account changing vaccination rates, mask wearing, and the relative prevalence of different SARS-CoV-2 variants. The model simulates daily contacts between trainees, trainers, and support staff during a ten-week training camp consisting of an initial two-week cocoon period with smaller training units of 60 trainees each, followed by the remaining eight weeks of camp when trainees are assigned to companies of 240 trainees each (Figure S1). Model calibration was conducted by fitting the model to the observed number of reported infections for training camps that began between October 2020 and April 2021. Model results show how outbreak size was associated with the number of introductions from newly arrived trainees or trainers and staff infected off base during training camp over the course of the pandemic. We then used the calibrated model to assess the impact of different control strategies including surveillance testing, mask-wearing, and vaccination mandates. We assessed the effectiveness of these strategies over the course of the pandemic as importation risk and population-level susceptibility changed considerably. Our results highlight the importance of addressing the broader community context when designing prevention and control efforts for high-risk settings.

## Results

### Infections in arriving trainees and correlations in observed infections

The observed portion of new military trainees testing positive by RT-PCR for SARS-CoV-2 infection on arrival at camp between the first week of October 2020 and the first week of October 2021 ranged from 0.6% to 10.7% (Figures 1A,2A). There was a significant correlation between the portion of trainees testing positive on arrival and the portion of trainees testing positive during the initial two-week cocoon period (r=0.47, p=0.016). There was also a significant correlation between the portion of trainees testing positive on arrival and the portion of trainees testing positive during the entire training camp (r=0.46, p=0.020), as well as a significant correlation between the portion of trainees testing positive during the initial cocoon period and the portion of trainees testing positive during the post-cocoon period (r=0.66, p=0.001; Figure 1B). Although the number of trainers and support staff infected off base during training camp was not reported, there was a significant correlation between incidence in the surrounding county during training camp and both the number of trainees testing positive on arrival (r=0.69, p<0.001), and the number of positive tests during the entire training camp (r=0.43, p=0.029; Figure 1C).

**Figure 1.**
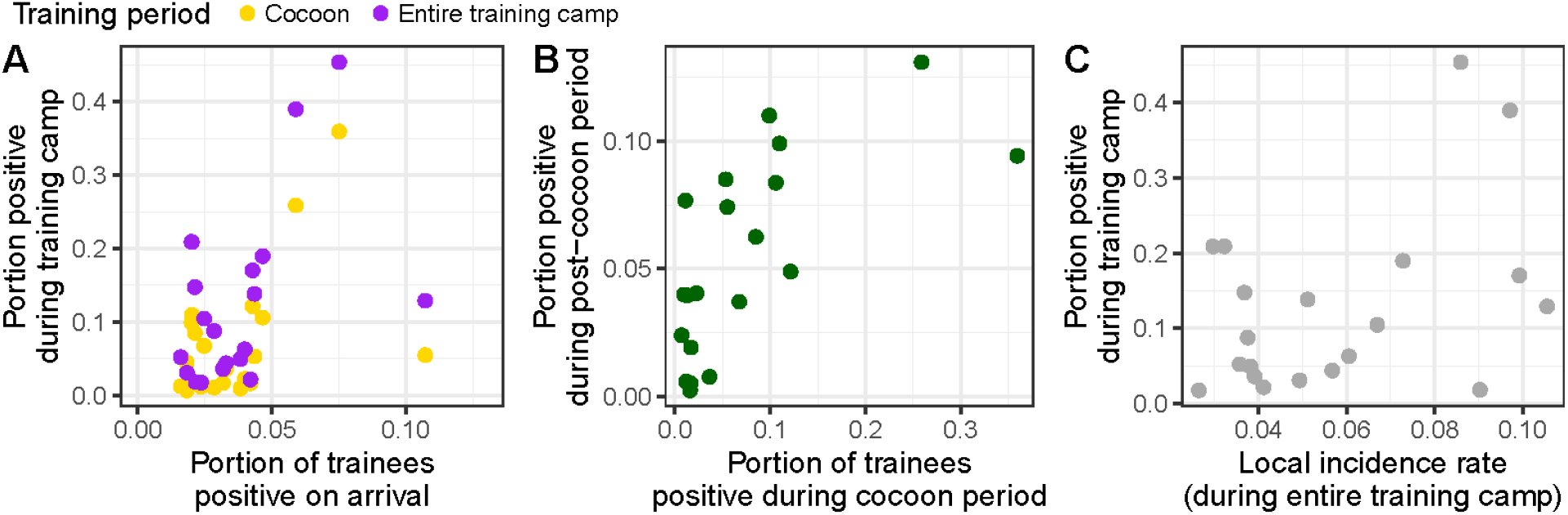
Correlations between observed SARS-CoV-2 positive tests. (A) Correlation between the number of new trainees testing positive upon arrival at training camp and the number of positive tests during the initial two-week cocoon period or the entire ten-week training camp. (B) Correlation between the number of positive tests during the cocoon and post-cocoon periods. (C) Correlation between incidence in the adjacent county during the ten-week training camp and the number of positive tests during training camp.

From October 2020 to October 2021, our median estimate of the portion of new arrivals expected to test positive ranged from 1.9% (95% prediction interval (PrI): 1.1-2.8%) on 10/10/20, to a peak of 8.8% (95% PrI: 7.1%-10.9%) on 01/16/2021, followed by a decline and then a second peak of 6.2% (95% PrI: 4.8-7.8%) on 09/04/2021 following the spread of the delta variant (Figure 2A). The 95% prediction interval of our estimates overlapped with the 95% confidence interval for the observed number of positive trainees on arrival in 35 of 44 weeks during this time period. The correspondence between observed and simulated positive trainees on arrival indicates that adjusted, national-level infection estimates provide a useful, independent prediction of the risk of importation into this setting. Simulating beyond our observation period, our model predicts a large omicron wave starting in late 2021 and peaking at 9.3% (95% PrI: 7.5-11.2%) on 01/22/2022. Following this peak, the portion of new trainees testing positive on arrival declined rapidly up to March 2022 and was forecasted to continue to decline to 0.8% (95% PrI: 0.4-1.5%) by 03/19/2022 based on CDC projections (*27*). The modeled fraction of trainers and support staff infected off base ranged from a low of 0.0% (95% PrI: 0.0-2.9%) for the training camp beginning on 05/01/2021 to a high of 10.0% (95% PrI: 4.0-15.0%) for the training camp beginning on 12/04/2021. Because all of the off-base infections over the 10-week training period are grouped by the camp start date in Figure 2E, these peaks occur earlier than the peaks in the number of infections in newly arrived trainees.

**Figure 2.**
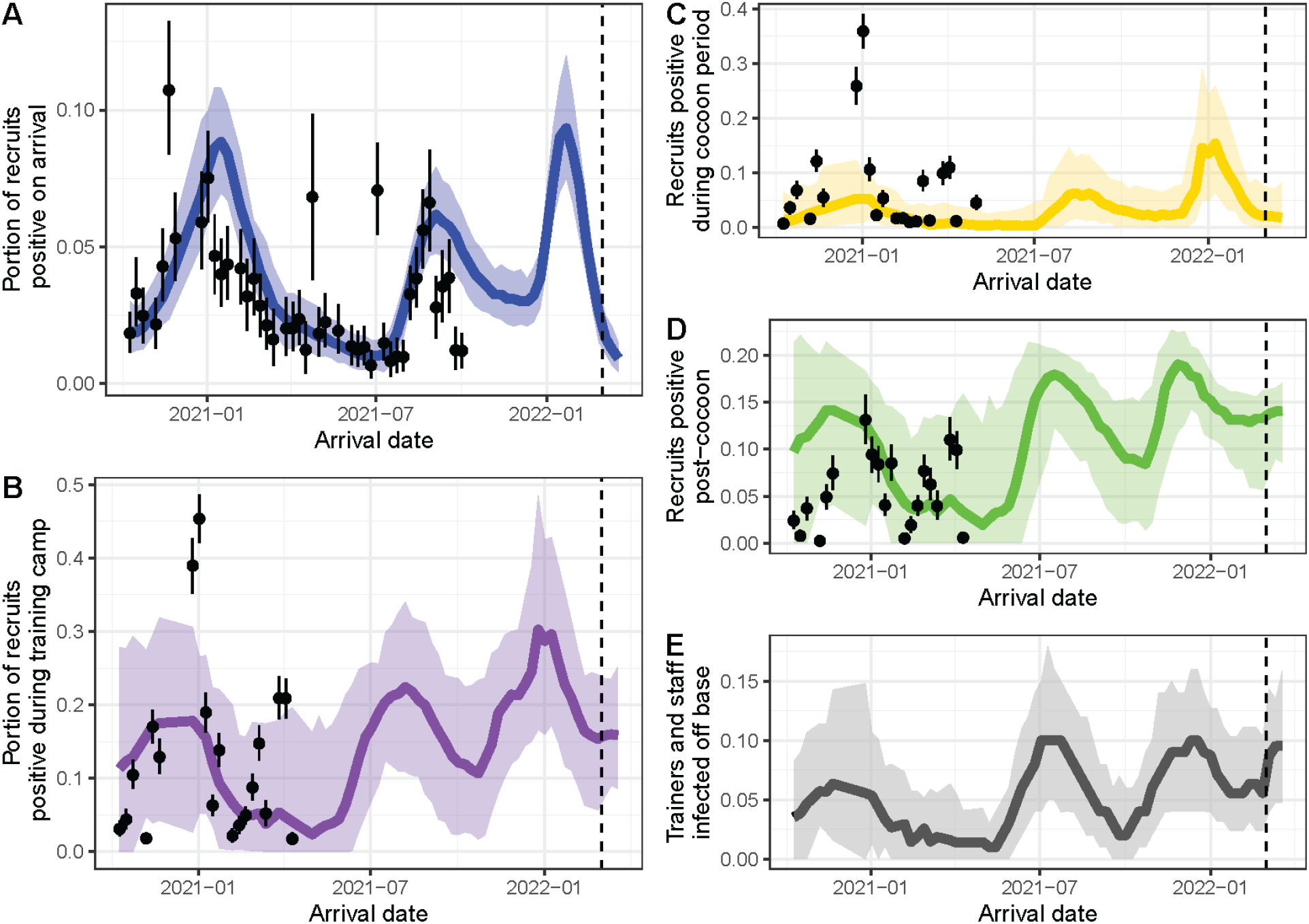
Portion of individuals infected before or during training camp. (A) Portion of new recruits testing positive upon arrival, (B) Portion of trainees testing positive during the initial cocoon period, (C) portion of trainees testing positive during the post-cocoon training period, and (D) portion of trainers and support staff infected off base during training camp. The red circles in (A-C) are the observed portion of positives for each time period. Dashed line represents the start of the period when estimates are based on forecasted cases from the CDC forecasting project (*27*).

### Basic reproduction number, R_0_, and training camp transmission patterns

The estimated basic reproduction number, R_0_, for the original SARS-CoV-2 during training camp was 6.5 (95% credible interval (CrI): 6.0-6.8). Based on estimates of their increased transmissibility (see Table S2), the resulting R_0_ values for the major variants of concern during the cocoon period were 8.4 (95% CrI: 7.7-8.8) for alpha, 9.0 (95% CrI: 8.3-9.4) for gamma, 12.9 (95% CrI: 11.8-13.5) for delta, and 18.6 (95% CrI: 17.1-19.5) for omicron. The estimated dispersion parameter for R_0_ was 4,466 (95% CrI: 501-10,000), indicating that there was not strong evidence for overdispersion of transmission in this setting. Overall, our model was able to recreate the observed outbreak size during the entire training camp, with the 95% prediction interval for the portion of recruits testing positive during training camp overlapping with 15 out of 20 observations (and 17 out of 20 of the 95% confidence intervals for the observations). The simulated portion of trainees testing positive during the cocoon period matched the observed number of trainees testing positive during this period well, with the exception of two large observed outbreaks at the end of 2020 (right after the holiday break when most trainees travel off base, a phenomenon not captured in our model) and three smaller outbreaks in March-April 2021 (Figure 2C). The predicted number of individuals testing positive during the post-cocoon period of training camp also matched observations well, although several larger outbreaks fell above the 95% range of the simulations in 2021 (Figure 2C).

The expected number of individuals testing positive in both the cocoon and post-cocoon periods of training camp exhibited peaks during the delta and omicron waves in the summer and winter of 2021 that largely followed the curve of trainees testing positive on arrival and the expected number of introductions from trainers and staff infected off base (Figure 2E). However, both of these peaks were larger than the peak in early 2021, despite the number of positive arrivals not being higher than during the early-2021 peak, due to the increased transmissibility of the delta and omicron variants relative to the original virus. In particular, the expected number of positive individuals during the post-cocoon period peaked at 24.3% (95% PrI: 14.8-28.0%) for the camp starting 12/04/2021, compared to only 4.8% (95% PrI: 1.2-9.2%) for the camp that started 01/02/2021. This peak in infections during training camp also occurred earlier than the peak of arrivals on 01/22/2022 (Figure 2A). Although the number of positive arrivals was forecasted to decline in February and March 2022, the number of trainees testing positive during the post-cocoon training period remained high, likely due to the high transmissibility of the omicron variant and the waning protection of individuals who were vaccinated or infected earlier in 2021.

### Association between importations, R_0_, and outbreak size

To examine the impact of importations and R_0_ on outbreak size, we compared simulated outcomes in training camps starting each week, as both importations and R_0_ varied over the course of the pandemic. However, because other factors also varied over the same timeframe (e.g., vaccination coverage, variants, levels of naturally-acquired immunity), their impact cannot be isolated, adding uncertainty to our estimates of the associations between importations or R_0_ and outbreak size. The number of infections during the cocoon period showed a positive correlation with the number of new trainees expected to be infectious upon arrival (based on the number infected in the two weeks prior to arrival) (Figure 3A), but there was no clear relationship between the number of potentially infectious new arrivals and the total number of individuals infected during the entire training camp (Figure 3B). However, there was a strong positive relationship between the predicted number of staff infected off base and the total number of individuals infected during training camp, although the relationship appears to reach a saturation point as the infection rate during camp approaches 100% (Figure 3C). The relationship between outbreak size (the portion of individuals infected during training camp) and the number of infectious arrivals, or staff infected off base, was similar across the entire period of study, but outbreak sizes were much larger per importation when the delta and omicron variants predominated (Figure S2). Outbreak size was also positively correlated with the mean R_0_ at the start of training camp, although this relationship was nonlinear, partially due to changes in the effective reproduction number (R_t_) over the course of the pandemic (Figure 4A). The effect of R_0_ on outbreak size persisted even when controlling for importation pressure, with outbreak size increasing as either R_0_ or the rate of introductions from off base increased (Figure 4B).

**Figure 3.**
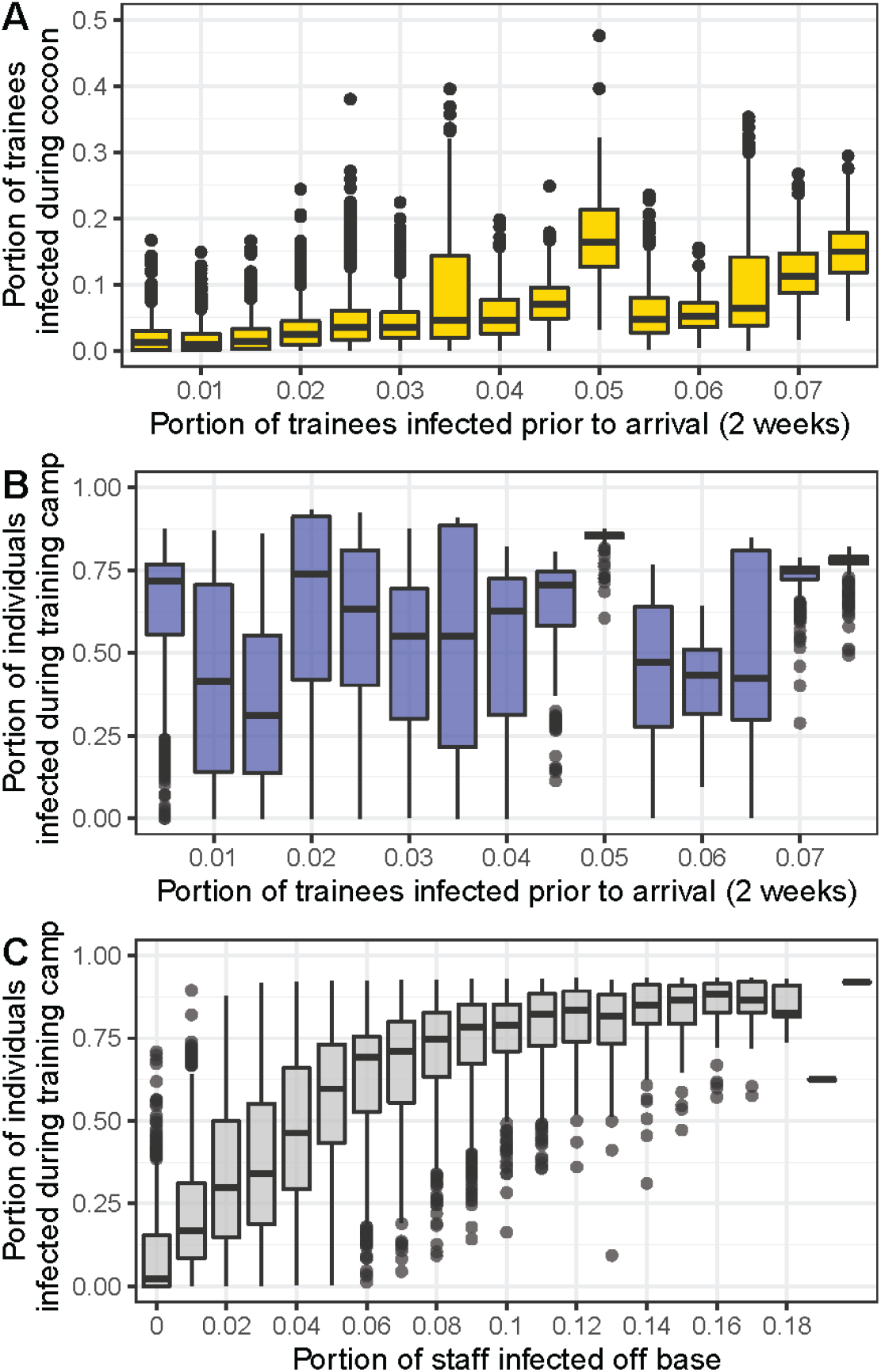
Effect of portion of trainees infectious upon arrival or portion of staff infected off base on outbreak size. (A) Portion of trainees infected during the cocoon period or (B) the portion of all individuals infected during training camp as a function of the predicted number of trainees who were infected in the two weeks prior to arrival. (C) Portion of all individuals infected during training camp as a function of the predicted number of staff who were infected off base.

**Figure 4.**
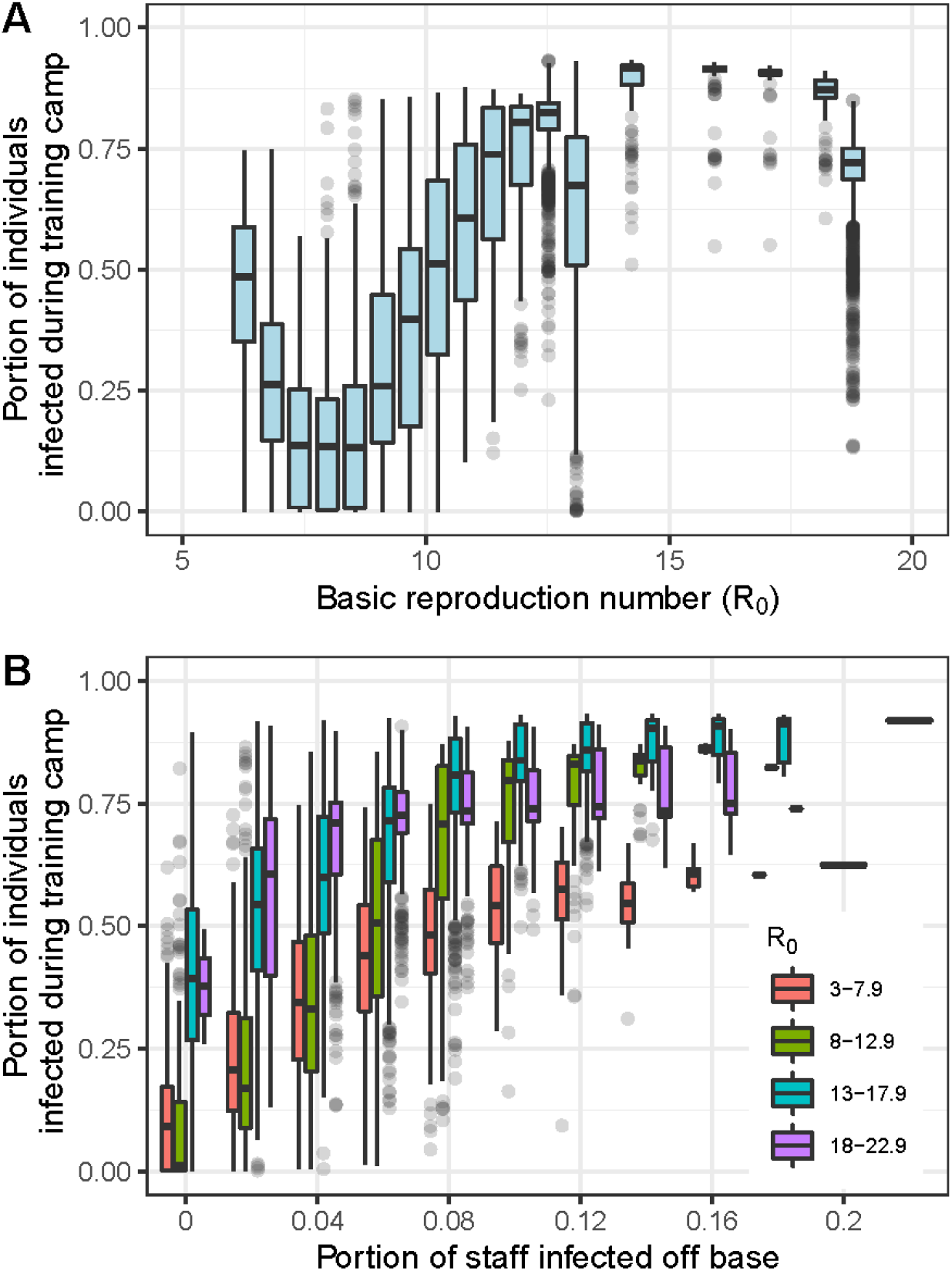
Effect of R_0_ on outbreak size. (A) Portion of all individuals infected during training camp as a function of the mean estimated basic reproduction number, R_0_, at the start of training camp (based on the relative proportions of different variants of concern currently circulating nationally). (B) Portion of all individuals infected during training camp as a function of the predicted number of staff who were infected off base and R_0_ at the start of training camp.

### Impact of existing interventions

Our model predicts that the percentage of new trainees testing positive during the cocoon period would have been noticeably higher without active surveillance testing on arrival (Figure 5A). For example, for the training camp starting on 01/02/2021, the percentage of trainees testing positive at the end of the cocoon period was projected to be 10.5% (95% PrI: 2.9-20.9%) with arrival testing, but 30.7% (95% PrI: 19.3-41.5%) without testing. The differences were even larger when the delta and omicron variants were dominant. At the peak of the delta wave on 08/21/2021, the predicted number of positive trainees was 15.1% (95% PrI: 4.5-27.8%) with arrival testing and 43.8% (95% PrI: 27.7-54.7%) without arrival testing. At the peak of the omicron wave on 01/08/2022, the predicted number of positive trainees were 26.0% (95% PrI: 13.2-43.7%) and 66.5% (95% PrI: 49.8-77.8%) with or without arrival testing, respectively. Over the whole simulation period, the median number of positive trainees during the cocoon period was 5.6% with arrival testing (95% PrI: 0.1-27.8%) and 20.1% (95% PrI: 2.4-65.0%) without arrival testing. The impact of arrival surveillance testing on the attack rate over the entire training period was not as large as the impact during the initial cocoon period (Figure 5B), because arrival testing did not reduce transmission in the post-cocoon period, and for some training periods the attack rate was higher in the post-cocoon period with arrival testing due to the smaller number of trainees who were infected during the cocoon period (Figure 5C). In contrast to the large impact of arrival testing on transmission during the cocoon period, model simulations suggest that surveillance testing at the end of the cocoon period had only a small impact on the percentage of trainees who subsequently tested positive during the remainder of the training period (Figure 5B,D). The median number of trainees testing positive during the post-cocoon training period was 9.8% (95% PrI: 0.0-24.1%) with testing and 11.6% (95% PrI: 0.0-25.7%) without active surveillance testing at the end of the cocoon period.

**Figure 5.**
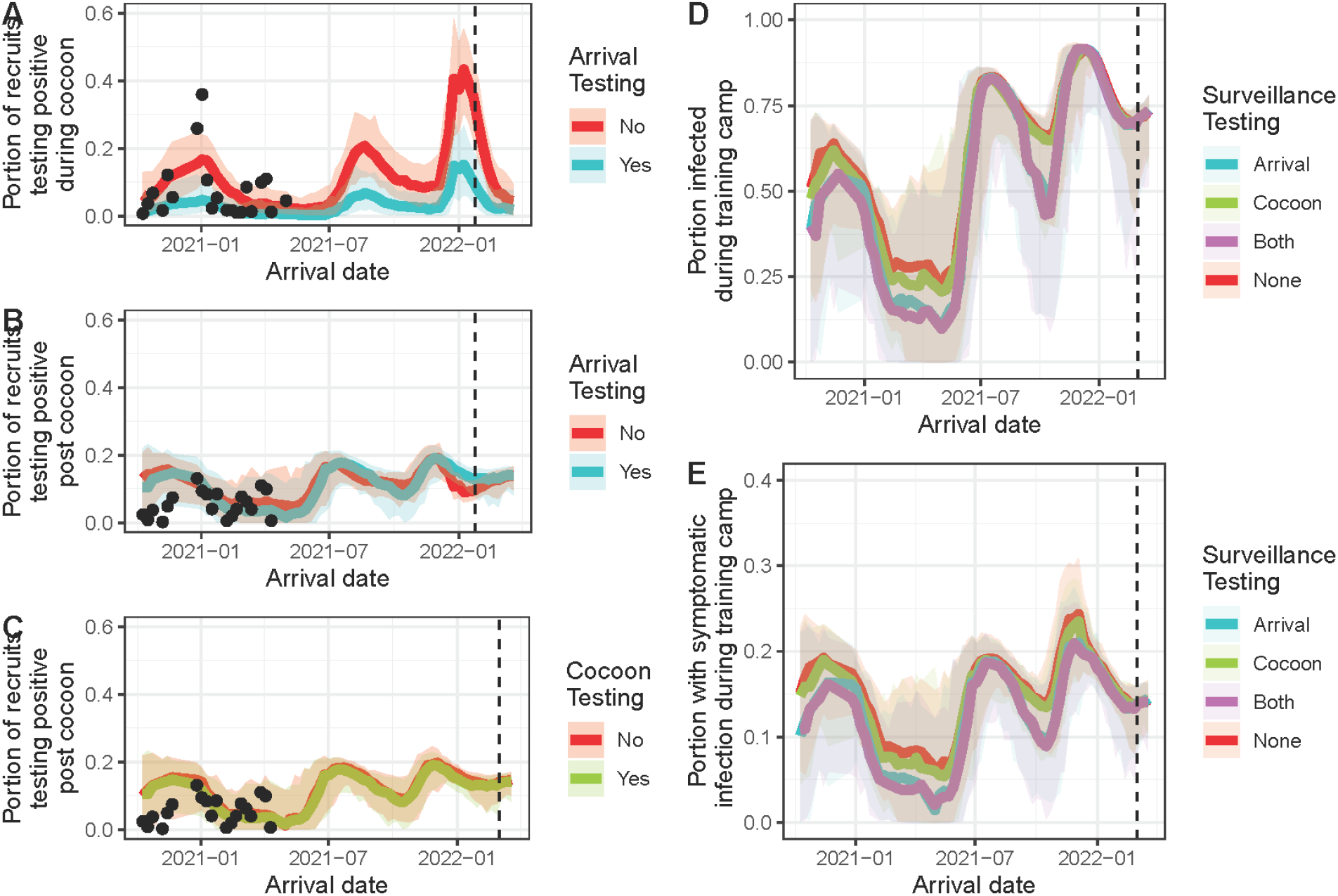
Impact of surveillance testing on outbreak size. (A) Portion of trainees (recruits) testing positive during the initial cocoon period with and without active surveillance testing upon arrival. (B) Portion of all individuals infected during training camp with and without active surveillance testing upon arrival. (C) Portion of trainees (recruits) testing positive during the post-cocoon training period with and without active surveillance testing upon arrival. (D) Portion of trainees (recruits) testing positive during the post-cocoon training period with and without active surveillance testing at the end of the cocoon period. The black circles in (A,C,D) are the observed portion of positive trainees for the appropriate time period. Dashed line represents the start of the period when estimates are based on forecasted cases from the CDC forecasting project (*27*).

Mask wearing also had a substantial impact on the number of individuals who were infected during training camp (Figures 6A,B). The median number of individuals infected during training camp was 44.5% (95% PrI: 0.2-94.4%) under our baseline scenario where compliance with masking guidelines varied over the course of the pandemic. When no masks were worn at all during training camp, a median of 57.2% (2.2-98.0%) of individuals were infected, and when no masks were worn and there were also no surveillance tests upon arrival or at the end of the cocoon period, 69.9% (95% PrI: 16.2-97.3%) were infected. Under an alternate scenario where mask-wearing compliance was 100% (except while sleeping or eating), only 31.1% (95% PrI: 0.2-90.3%) of individuals were infected during training camp. The voluntary vaccination of unvaccinated trainees upon arrival at training camp did not have a large impact on the number of individuals infected during training camp (Figures 6C,D). Compared to the baseline infection attack rate of 44.5% (95% PrI: 0.2-94.4%) with voluntary vaccination starting on 05/22/2021, the median infection attack rate was 45.7% (95% PrI: 0.4-96.4%) without voluntary vaccination upon arrival, and still 42.0% (95% PrI: 0.3-91.2%) even with a 100% uptake rate among unvaccinated trainees. The prediction intervals for the attack rates presented in this section are often quite wide because they incorporate both model stochasticity and variability in epidemiological conditions (e.g., variants, importation pressure, vaccination coverage) over the course of the pandemic.

**Figure 6.**
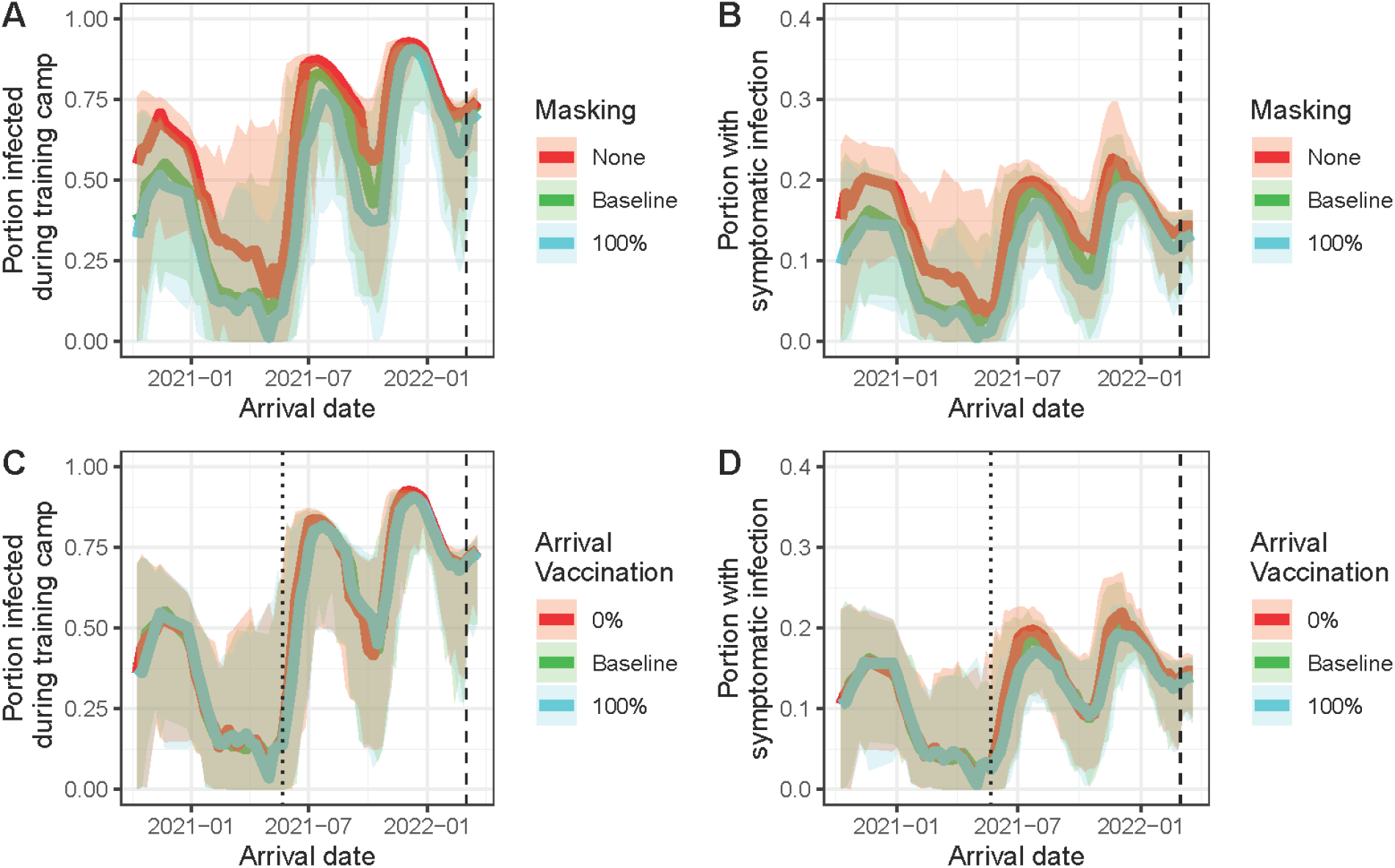
Impact of mask wearing or vaccination on outbreak size. (A) Impact of mask wearing on the portion of all individuals infected, and (B) portion of individuals with symptomatic infections during training camp. Baseline mask wearing rate varied over the course of the pandemic (see Methods). (C) Impact of vaccinating un-vaccinated trainees upon arrival at training camp on the portion of all individuals infected and (D) the portion of individuals with symptomatic infections during training camp. Baseline vaccination rate varied over the course of the pandemic (see Methods). Dotted line is the date for the start of vaccination requirement (05/22/2021).

### Impact of alternative intervention scenarios

In addition to assessing the impact of interventions that were implemented, we also examined the potential impact of several interventions that were not implemented: requiring vaccination of all trainers and staff by 05/22/2021 (with a booster administered on 12/01/2021), weekly or daily antigen testing of trainers and staff, or requiring that new trainees provide proof of full vaccination prior to the start of training camp beginning on 05/22/2021 (with a booster required after 12/01/2021). Because of the delayed rollout of the vaccine, here we focus on the impact of these interventions on training camps starting after 05/31/2021. Of the additional interventions, requiring daily antigen testing for trainers and staff had the largest impact on infection attack rates, reducing the number of individuals infected during training camp by 26.8% (95% CrI: 23.9-29.7%; t=18.5, p<0.001) and the number of symptomatic infections by 28.0% (95% CrI: 24.6-31.5%; t=16.3, p<0.001) compared to the default intervention scenario (Figure 7). Requiring new trainees to be fully vaccinated prior to arrival only reduced total infections by 7.1% (95% CrI: 5.1-9.2%; t=7.0, p<0.001), but reduced the number of symptomatic infections by 27.5% (95% CrI: 24.2-30.9%; t=16.7, p<0.001; Figure 7). Both of these interventions had a greater impact on symptomatic infections than masking, which was the most effective of the implemented interventions after 05/31/2021, with 100% masking reducing symptomatic infections by 12.8% (95% CrI: 10.9-14.8%; t=13.5, p<0.001) relative to the default masking scenario and by 21.1% (95% CrI: 17.9-24.5%; t=13.0, p<0.001) relative to a scenario with no masking (Figure 7). Requiring staff vaccination had a more modest impact, reducing infections by 6.5% (95% CrI: 4.6-8.5%; t=6.7, p<0.001) and symptomatic infections by 7.7% (95% CrI: 5.3-10.1%; ; t=6.4, p<0.001). However, combining all three additional interventions was highly effective against both total infections and symptomatic infections, reducing these by 47.0% (95% CrI: 41.4-52.5%; t= 17.0, p<0.001) and 59.0% (95% CrI: 52.8-65.2%; t= 19.2, p<0.001) respectively, relative to the default scenario with arrival screening, variable masking compliance, and voluntary vaccination upon arrival. Compared to a scenario with no interventions, combining arrival screening and moderate masking compliance with the three additional interventions reduced all infections by 53.2% (95% CrI: 47.2-59.2%; t=18.0, p<0.001) and symptomatic infections by 66.8% (95% CrI: 59.5-74.1%; t=18.5, p<0.001; Figure 7).

**Figure 7.**
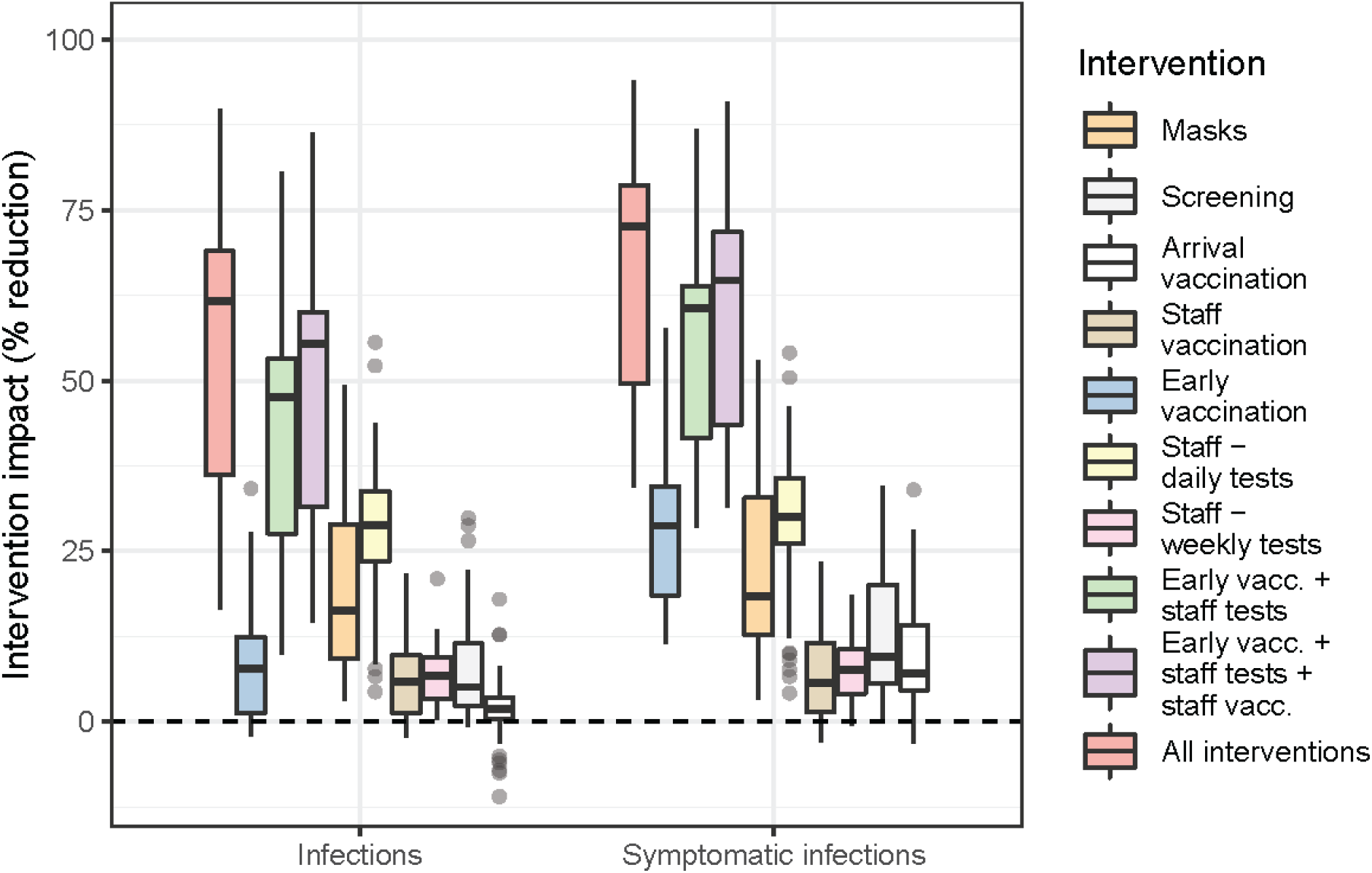
Impact of different intervention strategies on the total number of individuals infected and the number of symptomatic infections during training camp. All percent reductions are relative to the baseline scenario minus that particular intervention, except for the ‘All interventions’ scenario which is relative to a scenario with no interventions.

### Impact of importation pressure on intervention effectiveness

The impact of several individual interventions, and all interventions combined, weakened as the portion of new trainees infected in the two weeks prior to camp increased (Figure 8A). Fewer symptomatic infections were prevented by mask wearing (5.4 additional symptomatic infections per one percentage point increase (PI) in the infection attack rate, p=0.005,r^2^=0.18), requiring staff vaccination (2.7 additional symptomatic infections per PI, p=0.04,r^2^=0.10), daily staff antigen testing (7.3 additional symptomatic infections per PI, p<0.001, r^2^=0.37) and all interventions combined (9.0 additional symptomatic infections per PI, p=0.06,r^2^=0.08). Increases in the number of trainers and staff infected off base during training camp also altered the impact of several individual interventions, although it did not significantly reduce the impact of all interventions combined (Figure 8B). The only significant reduction in the impact on an intervention as off-base infections increased was for arrival screening, which prevented 4.1 fewer symptomatic infections per one percentage point increase in off-base infections (p<0.001, r^2^=0.24).

**Figure 8.**
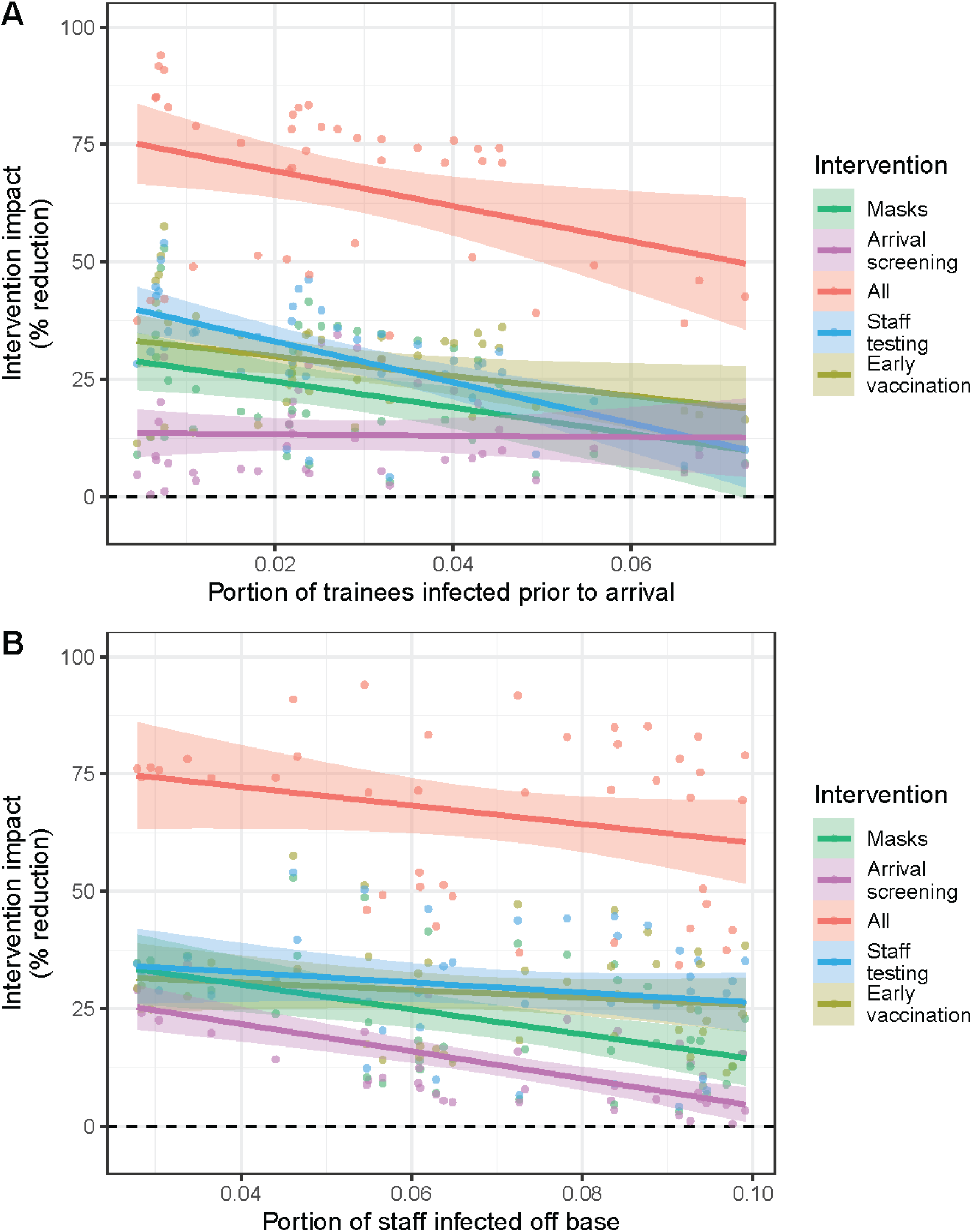
Effect of portion of trainees infectious upon arrival or portion of staff infected off base on impact of interventions. (A) The impact of several interventions as a function of the number of trainees who were infected in the two weeks prior to arrival. (B) The impact of several interventions as a function of the number of staff who were infected off base during training camp.

## Discussion

Our calibrated model results show that the risk of viral importation via newly arrived trainees, as well as off-base infections among trainers and support staff, varied considerably over the course of the SARS-CoV-2 pandemic. In addition, there was a positive correlation between the observed number of trainees testing positive on arrival and the observed fraction who tested positive during training camp. However, although both sources of introductions contributed to the amount of transmission occurring during training camp, the probability of an outbreak and final outbreak size were more strongly correlated with the number of trainers and support staff who were infected off base during the training period. The rate of viral introductions from off base was the strongest indicator of whether an outbreak would occur, but local conditions (as measured by the setting-specific R_0_) also influenced outbreak size. As more transmissible SARS-CoV-2 variants became dominant in this setting, outbreak sizes increased, even after accounting for the influence of introductions from off base. The combination of high importation rates and a high R_0_ value made control efforts difficult, and no single intervention was able to completely prevent outbreaks, particularly once the delta and omicron variants that had even higher R_0_ values became dominant. The most effective single intervention was daily antigen testing of trainers and support staff, which reduced the importation pressure from off-base infections. However, the impact of daily testing of staff, as well as the impact of several other interventions, was reduced when importation pressure was high from either newly arrived trainees or from off-base infections among staff. This suggests that preventing SARS-CoV-2 outbreaks in institutional settings requires tailoring multiple interventions that respond to importation pressures from the broader community, in addition to controlling transmission within the setting.

Despite the modest correlation between the observed number of positive trainees upon arrival at training camp and the number of trainees testing positive during training camp, our model suggests that SARS-CoV-2 introductions from trainers and support staff infected off base during training camp had a much stronger influence on outbreak risk and size. This result aligns with our previous research, which also highlighted the importance of off-base infections among trainers and support staff in driving outbreak risk (*8*). Introductions from off-base infections are important drivers of outbreak risk because they can seed new outbreaks at any time during training camp, and support staff who interact with trainees from different companies can seed outbreaks in multiple cocoons or companies. Other institutional settings face similar issues with importation pressures from certain community members who have contact with the broader community on a regular basis and can therefore introduce the virus at any time: prison staff and visitors (*28*), nursing home staff and visitors (*18*), or off-campus infections among college and university students following pre-matriculation screening (*12, 29, 30*). These individuals are also of particular importance for broader prevention efforts because, during an outbreak within an institutional setting, they can also spread the virus in the opposite direction back into the broader community (*1, 31*).

Of the interventions that were implemented during basic training in 2020 and 2021, our model suggests that mask wearing was the most effective at reducing the number of both symptomatic and total infections. However, the effectiveness of mask wearing did decrease as importation pressure from newly arrived trainees or staff infected off base increased, and at the highest observed rates of positivity among newly arrived trainees, arrival PCR screening was equally as effective as mask wearing at reducing infections during training camp. Mask wearing was also not effective enough to completely prevent outbreaks except when importations were low over the entire training period. The effectiveness of mask wearing is reduced in this setting by the communal sleeping quarters and communal dining. In addition, the most effective N95 or KN95 masks were not required, and anecdotal evidence suggests that compliance was well below 100% during certain periods of the pandemic. However, increasing masking compliance to 100% in our model only led to modest reductions in outbreak size during most periods of the pandemic. PCR screening of all newly arrived trainees also lowered the number of infections in training camp and maintained its effectiveness even when positivity rates among incoming trainees were at their highest. However, its impact on transmission was limited to the first two weeks of training camp (the cocoon period), and its impact was minimal when introductions via off-base infections of trainers and staff were high. The final implemented intervention, offering voluntary vaccination of unvaccinated trainees upon arrival, had almost no impact on outbreak size and did not reduce imported infections due to the timing of the intervention.

Our model results indicate that several interventions that were not implemented might have had a larger impact on outbreak risk—and outbreak size—than those interventions that were implemented (surveillance testing, masking, and voluntary vaccination for new trainees upon arrival). The intervention with the largest average impact over the course of the pandemic was daily antigen testing of trainers and support staff. The large impact of this intervention highlights the importance of preventing, or at least reducing, the number of viral introductions via off-base infections. The impact of testing trainers and staff did decline as importation pressure from newly arrived trainees increased. As a result, estimating importation pressure from regional or national incidence patterns may be an important component of SARS-CoV-2 prevention and control efforts in this and similar institutional settings. Knowledge of importation pressure can be used to tailor the prevention strategy, ideally by implementing multiple interventions that target different sources of introduction into the community (e.g., screening both incoming trainees upon arrival and testing trainers and support staff who live off base on a regular basis). Unlike daily antigen testing, weekly surveillance testing of trainers and support staff via PCR or antigen tests had minimal impact on outbreak size, likely due to the short generation interval of SARS-CoV-2 and the quick rise in infectiousness within a few days of infection (*32*–*35*). Although daily antigen testing of staff had the largest impact on infections, it is likely not the most cost effective or feasible intervention due to the large number of tests that would be required on a daily basis. Each training cohort is supported by up to 120 trainers and staff, and there are multiple training cohorts on base on any given day.

Requiring all new military recruits to be fully vaccinated prior to arrival at training camp had a smaller impact on outbreak size (total infections) than daily testing of staff, but had a similar impact on the number of symptomatic infections. In addition, early vaccination was more effective than testing staff when the importation pressure from new trainees was predicted to be high based on national incidence patterns. In this scenario, staff testing is less effective because it does not catch introductions via trainees, while vaccination of trainees prior to arrival both reduces the probability that they will be infectious on arrival (lowering importation pressure) and reduces transmission during training camp (particularly if asymptomatic infections are less infectious). In contrast to the impact of requiring vaccination prior to arrival, voluntary vaccination of trainees upon arrival had almost no effect on outbreak risk and only a small impact on the number of symptomatic infections during training camp. Post-arrival vaccination does not reduce SARS-CoV-2 introductions from arriving trainees, and full protection is only established about halfway through the ten-week training period, limiting the impact on transmission within the camp. Requiring all staff to be vaccinated by 05/31/2021 (and receive a booster dose by 12/01/2021) had a smaller impact on outbreak size than daily testing of staff or pre-arrival vaccination of trainees due to the rapid decline in protection from infection provided by vaccination.

Combining daily antigen testing of staff with a vaccine requirement for new trainees prior to arrival had an additive impact on reductions in both the total number of infections and symptomatic infections during training camp. Screening new trainees upon arrival, adding a vaccine requirement for staff, or requiring masking all further reduced outbreak size when added either individually or as a group. The continued impact of additional interventions showcases the importance of combining multiple interventions in institutional settings with high R_0_, where no currently available intervention is capable of entirely preventing transmission by itself (*36*–*38*). While all interventions combined reduced symptomatic infections by ∼75% when importation pressure from either new arrivals or staff living off base was low, an increase in viral introductions from either group lowered the overall impact. When importation pressure from newly arrived trainees was at its highest during the pandemic, all interventions combined only prevented about 50% of symptomatic infections, partly because the impact of daily antigen testing of staff was greatly reduced. This highlights the importance of understanding the current source of introductions when implementing intervention strategies, and suggests that at times of high importation risk additional interventions, such as stronger contact tracing and quarantine policies, might be necessary to prevent outbreaks in this or similar institutional settings. One caveat regarding these impact estimates is that they are for training camps that started after vaccination became routine (05/31/2021), and therefore, they mostly reflect periods when the highly transmissible delta and omicron variants were dominant. Outbreak sizes from the first year of the pandemic suggest that even without vaccination, a combination of surveillance testing, masking, and social distancing could limit outbreaks except when importation pressure was at its highest during the winter of 2020-2021.

Although viral introductions via off-base infections among trainers and staff were a major driver of outbreak size, transmission dynamics within training camp also played a major role, with higher R_0_ values leading to larger outbreak sizes at a given off-base infection rate. Our estimate of R_0_ for the original SARS-CoV-2 strain in this setting was higher than estimates from broader community settings (*39*–*41*), but similar to some estimates from high-risk settings like cruise ships (*2, 42*), as well as previous estimates from two outbreaks at other military bases (*8*). The reported number of positive tests during training camp also indicate frequent within-camp transmission, although the extent of these outbreaks is hard to determine because there was no active surveillance after the initial two-week cocoon period and only symptomatic individuals were tested and isolated. This evidence indicates that in addition to interventions intended to prevent introductions, such as screening new trainees upon arrival or daily testing of staff who live off base, control efforts that limit transmission within training camp remain an essential tool to limiting outbreaks, particularly as new variants with increased transmissibility have emerged. While vaccination remains a critical tool for preventing severe disease and hospitalization, its protection against infection is modest and declines rapidly, particularly for the omicron variants that are now dominant (*43, 44*).

By estimating the infection histories of new military recruits from national incidence patterns, our model’s predictions closely matched the number of new trainees testing positive upon arrival at training camp in 2020 and 2021. This result suggests that publicly available data can be used to assess population-level infection patterns and importation pressure into similar institutional settings. Based on knowledge about the demographic makeup of the population of military recruits (mostly 18-24 year olds) we adjusted infection estimates based on age-specific incidence patterns and geographically-weighted state-level estimates based on historical recruiting patterns. Other institutional settings, such as colleges and universities or summer camps, will have different, but potentially still identifiable, demographic and geographic compositions and could adjust infection estimates accordingly. Forecasting the importation pressure from the broader community is important for prevention and control efforts because our results show that even though arrival screening is relatively effective, it will still miss very recent infections (*45, 46*). While overall our predictions of positivity among newly arrived trainees closely matched the observed data, there were three weeks when the observed rates of positive infection were substantially higher than our predictions. One possibility for these outliers is that some trainees were infected during shared transportation from a processing center to training camp. Observed positive rates also declined more rapidly than our predictions following epidemic peaks in January and September of 2021, even though peak predictions closely matched observed positive rates. This suggests that actual infections may have fallen more rapidly than estimated by Chitwood et al. (*47*), at least among the new military trainee cohort.

### Limitations

One limitation of our analysis was that while we estimated off-base infections among trainers and support staff, we did not have any observational data with which to validate those estimates. If the infection risk among military staff was different from the estimated risk among adults in Richland County, SC due to differences in their community contact patterns, then these estimates might not accurately reflect the importation pressure from this source. Another limitation of our model is that we did not have detailed information on contact patterns among trainees or between trainees and staff during training, and therefore contacts within a training unit were randomly assigned and assumed to be of equal duration and strength. Our model therefore is not capable of capturing the impact of contact heterogeneity among trainees on transmission dynamics or the potential role of superspreaders due to the constraints on the number of contacts each agent can have. Given the importance of transmission from trainers and staff to trainees in our model, it would also be useful to assess whether contacts between trainees and staff actually have the same transmission potential as trainee-trainee contacts. Finally, we did not consider the cost-effectiveness or the feasibility of implementing the different interventions included in our analysis. Logistical constraints could outweigh the epidemiological effectiveness of certain interventions depending on the setting.

### Conclusion

Our model highlights how SARS-CoV-2 transmission patterns in the outside community affect outbreak risk in institutional settings similar to the military training camp considered in this study. Outbreak size was highly correlated with the importation pressure from off-base infections among staff, and our model indicates that the single most effective intervention would have been daily antigen screening of trainers and staff living off base. We also found that the effectiveness of some interventions (e.g., arrival screening of new trainees and masking requirements) decreased as importations from off-base infections of staff increased, while the effectiveness of other interventions (e.g., daily screening of staff, vaccination of staff, masking requirements) decreased as importations from newly arrived trainees increased. These influences of viral introduction rates on intervention effectiveness highlight the importance of tailoring multiple-intervention strategies in institutional settings to address both sources of introduction and local transmission. The correspondence between our estimated infection rates and observed positivity rates among newly arrived trainees also indicates that publicly-available incidence data can be used to predict importation pressure and outbreak risk in real-time, and therefore could inform outbreak prevention and control efforts in institutional settings such as military bases, prisons, schools, and assisted-living facilities.

## Methods

To examine SARS-CoV-2 transmission dynamics during military basic training, we developed an agent-based model that captures key features of this institutional setting. The model tracks a single cohort of new military recruits (trainees) through the 10-week training period and simulates daily contacts between individual trainees, their trainers, and support staff (Figure S1). Full details of the original model are described in España et al. (*8*). For this analysis we updated the original model by incorporating temporal dynamics in the importation risk from outside of the military base, vaccination, waning of immunity from natural infection and vaccination, and the introduction of new SARS-CoV-2 variants of concern. The model was implemented in the R programming language (*48*). Model code, as well as R code to calibrate the model and analyze model results are available at https://github.com/confunguido/basic_training_dynamics.

### Structure of agents and contacts

The model is designed to simulate the daily interactions of new military recruits (trainees), along with their trainers and support staff during the initial ten-week training camp. Individual agents in our model are classified as either trainees, military trainers, or other support staff (Figure S1). We assumed that all new trainees arrive as a single cohort at the beginning of training camp each week and are initially assigned to cocoons containing 60 trainees each, with two trainers assigned to each cocoon. During the initial two-week cocoon (quarantine) period there is no contact between trainees or trainers in different cocoons, but individuals also have contacts with support staff (dining, equipment, etc.) who interact with multiple cocoons. At the end of the two-week cocoon or quarantine period, the trainees, trainers, and support staff from each cocoon are pooled into larger companies with a maximum of 240 trainees each. The default weekly cohort size is 1200 trainees (20 initial cocoons and 5 companies), but most training camps in 2020 and 2021 had smaller cohorts, and a few had over 1200 (a maximum of 1254). We assume that trainees do not leave base during training camp, but that trainers and support staff leave the military base each evening and can therefore become infected in the outside community.

Prior to simulating transmission dynamics during training camp, the model was first initialized by simulating an infection and vaccination history for each individual based on national incidence and vaccination data (see below). We then constructed a contact network for the entirety of training camp, with trainee-trainee and trainer-trainee contacts restricted to within-cocoon contacts during the initial two-week cocoon period, and to within-company contacts during the post-cocoon period. During the cocoon period, trainees were randomly assigned contacts within their cocoon using a binomial distribution with a probability of 0.2 (a mean of 12 trainee contacts per individual for cocoons of 60 trainees). During the post-cocoon period, trainees were randomly assigned contacts within their company using a binomial distribution with a probability of 0.1 (a mean of 24 trainee contacts per individual for companies of 240 trainees). Trainers were assumed to have contact with all trainees in their assigned cocoon, which continued in the post-cocoon period when cocoons had been merged into larger companies. Support staff were randomly assigned contacts with trainees from any cocoon or company. We assumed that the mean number of contacts per staff member was 20. The number of contacts for each staff member was drawn from a multinomial distribution, with the size equal to the number of staff multiplied by the mean number of contacts, and the contact probability drawn from a Beta (1,1) distribution.

### SARS-CoV-2 infection and transmission

SARS-CoV-2 introductions in our simulations occurred either at the start of the simulation via newly arrived infected trainees, or throughout training camp when trainers and support staff were infected off base (see below for description of initial importations). During the training camp, the probability of an infected agent transmitting the virus to a susceptible contact was calculated based on the basic reproduction number (R_0_), the time since infection, and the generation interval distribution, which we approximated with a Weibull distribution (see Table S1 for generation interval parameters). The probability of a symptomatic individual infecting each immunologically-naïve (completely susceptible) contact on day *t* of their infection is

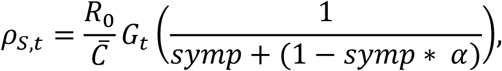

where 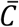 is the average number of contacts, *G*_*t*_ is the generation distribution on day *t, symp* is the probability of an infection being symptomatic, and *α* is the reduction in infectiousness for asymptomatic infections. For asymptomatic individuals, the probability of transmission to each immunologically-naïve contact on day *t* is reduced by *α, ρ*_*A,t*_ = *ρ*_*s,t*_*α*. Therefore, R_0_ is equivalent to the sum of daily transmission probabilities across all days of the generation distribution multiplied by the average number of contacts, 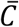, and weighted by the probability that an individual develops a symptomatic infection, 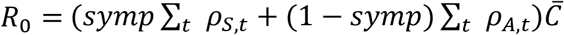. The probability of transmission from an infectious individual (*ρ*_*s,t*_ or *ρ*_*A,t*_) would be reduced if a susceptible contact had some partial immunity to infection from a prior infection or vaccination. The probability of transmission would also be reduced if either the infectious individual or their contact (or both) wore a facemask (see Table S1 for mask efficacy).

Symptomatic infections manifested according to an incubation period drawn from a discretized gamma distribution (*45*) and lasted for a certain number of days drawn from a Poisson distribution (*49*). We assume that trainees are tested via PCR on the day that they become symptomatic. Trainees that tested positive were placed in a communal sick bay and remained there for a minimum of ten days or until their symptoms resolved. Trainers and support staff testing positive isolated at home off base for a minimum of ten days or until their symptoms were resolved.

### Waning of natural and vaccine-derived immunity

Following an infection, we assume that an individual is completely immune to reinfection by all variants for the first 30 days due to the immune response to the primary infection. Individuals remained immune to reinfection from the same variant from day 30 to day 90 post-infection, after which immunity waned exponentially according to estimates from Townshend et al. (*50*). Cross-protection against other variants was modeled as a reduction in immunity relative to immune protection against the infecting variant and was assumed to wane at the same rate. Variant-specific cross-protection estimates are provided in Table S2. The first, second, and booster vaccine doses were assumed to provide protection against infection 14 days after the dose was administered. Initial protection against infection from the original SARS-CoV-2 virus was 36.8%, 77.5%, and 77.5% respectively for these three doses (*51*), with lower levels of protection for several variants of concern (Table S1 and S2). Protection against infection via vaccination waned according to a logistic equation fit to the estimates of protection at 1-6 months (*51*).

The increased transmissibility of the alpha, gamma, and delta variants relative to the transmissibility of the original SARS-CoV-2 strain were obtained from published estimates (Table S2). In the absence of a reliable estimate of the increased transmissibility of the omicron variant that isolated the separate impacts of increased transmissibility and immune escape on its relative fitness, we estimated the relative transmissibility based on the results of a modeling study that examined plausible ranges of transmissibility and immune escape (*52*). Assuming that protection against an omicron infection from a previous SARS-CoV-2 infection was 56% (*53*), we selected the most plausible relative transmissibility according to the estimates of (*52*). For each variant, the relative transmissibility was treated as a multiplier on the R_0_ of the original virus.

### Model initialization

Prior to the start of each training camp, we first simulated an infection and vaccination history for each individual agent. The prior infection history and vaccination status of individuals varied significantly over the course of the pandemic (*13*), and likely had a major impact on transmission dynamics in the military training setting. Therefore, for each cohort we simulated prior infection histories using estimates of daily infections at the state-level from CovidEstim (*47*). These state-level infection estimates were age-adjusted using national-level age-specific case data from the CDC (https://data.cdc.gov/Case-Surveillance/COVID-19-Case-Surveillance-Public-Use-Data/vbim-akqf) by multiplying the proportion of the population infected on a particular day by the weekly relative risk of infection for the 20–29-year-old age group. The age-adjusted, state-level infection estimates were then aggregated to the national-level with each state weighted by the percentage of military recruits from that state in 2018 (https://www.cfr.org/backgrounder/demographics-us-military). The infection estimates were provided as median, 2.5%, and 97.5% confidence intervals, from which we calculated a daily mean and standard deviation. For each trainee, we then used the daily infection estimates to generate an infection history prior to their arrival at training camp. The probability that an individual was infected by the original SARS-CoV-2 virus or one of the alpha, gamma, delta, or omicron variants was estimated using weekly sequence data from GISAID and Covariants.org (*54*). The weekly proportion of sequences for each variant at the state-level were aggregated to the national level using the same geographical weighting as the infection estimates. The vaccination history for each arriving trainee was estimated in a similar manner using state-level daily estimates of the number of 18–24-year-olds receiving their first, second, and booster doses provided by CDC (*55*). We also generated infection and vaccination histories for trainers and support staff using the infection and vaccination estimates among individuals 18 or older for Richland County, SC where Fort Jackson is located. The potential infectiousness of newly arrived trainees at the start of training camp was determined by their simulated infection histories and the generation interval (Table S1). In addition to importations via newly arrived trainees, the virus can also be imported into the training camp by trainers and staff who live off base and return home every evening. The daily probability of one of these individuals being infected off base was estimated using the same daily CovidEstim infection estimates for Richland County, SC that were used to simulate pre-camp infection histories for trainers and staff.

### Baseline scenario

Surveillance testing via PCR was conducted for all new trainees when they arrived at training camp, and again at the end of the initial two-week quarantine period before cocoons were merged to form the larger companies. Any individuals testing positive during one of these tests were placed in a common isolation bay. During the remainder of training camp, testing and isolation of positives was only performed for symptomatic individuals. A detailed description of the model assumptions regarding test specificity and sensitivity are provided in España et al. (*8*). A mask-wearing requirement for trainees, trainers, and staff in camp has been in place throughout the pandemic per CDC guidelines for high-risk settings (*56*), except for when trainees were eating, sleeping, or attending to personal hygiene. However, anecdotal evidence suggests that compliance has not been universal, particularly during periods of low transmission nationally. Therefore, we assumed that masking compliance followed the daily reported mask wearing rate for South Carolina as surveyed by Delphi and reported in Covidcast (*57*), with an adjustment of 2/3 of this value to account for periods when trainees were indoors but not required to wear masks (sleeping, etc.). Starting the week of 05/22/2021, all new trainees who were unvaccinated or incompletely vaccinated were offered a two-dose vaccine regimen upon arrival. While weekly data on vaccine uptake at arrival was not available, based on anecdotal evidence we assumed that vaccine uptake upon arrival among unvaccinated trainees was 25% from 05/22/2021 to 06/30/2021, 40% from 07/01/2021 to 12/26/2021, and 60% after 12/26/2021. We assumed that the first dose was administered on day 3 of training camp, with the second dose administered 14 days after the first dose.

## Model calibration

We estimated the basic reproduction number (R_0_) for the original SARS-CoV-2 virus in this military training setting by calibrating the model to surveillance data from a U.S. Army training base. Between October 2020 and April 2021, all trainees were tested via PCR on arrival and at the end of the two-week quarantine period. In addition, we also obtained data for the same dates on the number of symptomatic individuals who tested positive via PCR during the remainder of training camp. The arrival test data was not used for model fitting, but was instead used to validate our estimates of the expected number of infections in newly arrived trainees based on national incidence. Initial estimation of R_0_ during training camp was done by running the model and assuming that the number of contacts infected by an infectious agent was approximated by a beta-binomial distribution with a dispersion parameter, *k*. We ran 4000 different parameter combinations using a sobol sequence with R_0_ ranging from 1-30 and the dispersion parameter, *k*, ranging from 10^-6 to 10^4. For each parameter combination we ran 200 simulations to capture parameter uncertainty and model stochasticity. The log likelihood for each simulation was estimated by summing the separate log likelihoods of the number of positives during the cocoon period and the post-cocoon training period using a beta-binomial distribution for each period. We then fit a generalized additive model with a Gaussian process smoothing term to produce a joint posterior distribution for R_0_ and the dispersion parameter.

## Analyses

### Model behavior under baseline scenario

Following calibration of the model and the estimation of R_0_, we simulated training camps starting on a weekly basis from October 10, 2020 through March 19, 2022. For training camps starting between 10/10/2020 and 04/10/2021 we used the reported cohort size, which varied from 631 to 1254. For all subsequent training camps, we assumed that there were 1200 trainees, along with 40 trainers and 60 support staff. We performed 200 replicate simulations for each week and examined the time course of the outbreak for each replicate, tracking the number of agents who were infected (including off base infections for trainers and support staff) during the initial two-week cocoon period and during the subsequent eight weeks of training camp. We ran these model projections through 03/02/2022 using the daily infection estimates from CovidEstim (*47*) and the variant data from Covariants.org (*54*). We then extended the model projections for an additional three weeks using the forecasted state-level case data from the 02/28/2022 four-week ahead forecast from the COVID Forecasting project (*27*). Weekly forecasted case numbers were converted to infection estimates by assuming that the probability of an infection being reported during the forecasted period would equal the mean reporting rate over the course of the pandemic, and then adjusting for the lag between the date of infection and the date a case would be reported.

### Assessing the potential impact of different interventions

To examine the importance of the surveillance testing, mask wearing, and voluntary vaccination of trainees on arrival we ran counterfactual scenarios with one or more of these interventions removed. We also examined the potential impact of several potential interventions that were not implemented. These additional interventions were enforcing 100% masking compliance, surveillance testing of trainers and staff, requiring vaccination of new trainees starting on 05/22/2021, and requiring vaccination of support staff and trainers starting on 05/22/2021 (with required booster shots also administered on 12/01/2021). Three different surveillance testing regimes were considered for staff: weekly testing via PCR, weekly antigen testing, or daily antigen testing. In addition to examining the impact of requiring all unvaccinated trainees to receive a full vaccine course on arrival (rather than arrival vaccination being voluntary), we also examined the impact of requiring trainees to be fully vaccinated before arriving at the beginning of training camp. For this scenario, we assumed that all trainees who were not already vaccinated 35 days prior to the start of training camp would receive their first and second vaccine doses 35 and 14 days prior to arrival respectively, so that they would have the maximum vaccine-derived protection upon arrival. For this pre-arrival vaccination scenario we also assumed that all trainees would receive a booster dose 14 days prior to arrival for all training camps starting after 12/01/2021. In addition to simulating each of these additional intervention scenarios individually for each weekly training camp, we also examined the impact of combining the staff surveillance, pre-arrival vaccination, and/or staff vaccination interventions.

The impact of each intervention was measured by comparing the mean number of infections or symptomatic infections from 200 simulations with and without that intervention. Statistical significance of intervention impact was assessed using a two-sample Student’s t-test. To permit comparison between all intervention types, statistical tests were restricted to the impact of interventions during training camps starting after 05/31/2021 following the implementation of vaccination requirements. To determine whether the impact of each intervention was influenced by the number of viral introductions from off base, we used linear regression models with mean difference in symptomatic infections with and without that intervention as the response variable and either (a) number of new trainees estimated to have been infected in the two weeks prior to arrival, or (b) the number of trainers and staff infected off base during training camp as the explanatory variable.

## Author Contributions

Conceptualization: SMM, TAP, TLH, KM, PTS, RMG

Methodology: SMM, GE, TAP, RMG, TLH, PTS, SP

Software: SMM, GE

Formal Analysis: SMM

Visualization: SMM

Validation: SMM

Data acquisition, curation, analysis: MEH, JBJ

Interpretation: RMG, PTS

Supervision: TAP, KM, PTS, RMG

Project administration: TAP, KM, SH

Funding acquisition: PTS, SP

Writing—original draft: SMM

Writing—review & editing: SMM, TAP, GE, KM, MEH, JBJ, RMG, SH, SP, RMG

## Data and materials availability

For questions related to the SARS-CoV-2 testing data used for model calibration, contact the Fort Jackson Public Affairs Office at. All publicly available data and all code used in this analysis are available at https://github.com/confunguido/prioritizing_interventions_basic_training

## Supplementary materials

**Table S1. Model parameters**.

**Table S2. Variant-specific model parameters**. Relative transmissibility is transmissibility of a variant relative to the original SARS-CoV-2 strain. Relative cross-protection is the amount of cross-protection against a particular variant that is provided by a past infection with a different variant, relative to the level of protection provided by a same-variant infection. Vaccine protection is the level of protection against infection by a particular variant relative to vaccine-derived protection against infection by the original SARS-CoV-2 strain.

**Figure S1. Model schematic**. Trainees arrive at the beginning of training camp (blue), progress to cocoons of 60 trainees each for 14 days (yellow), and then progress to companies of 240 trainees each for 56 days (green). Trainees have contact with other trainees in their cocoon or company, with trainers (brown) assigned to their unit (two per cocoon, eight per company), and with support staff (gray). Trainees who test positive following arrival testing or presentation with symptoms are placed in the sick bay (red) for ten days (or until symptoms resolve) before returning to their unit. Trainers and support staff who test positive following presentation with symptoms isolate from home for a minimum of ten days. All processes in the model are defined on a daily time step.

**Figure S2. Effect of portion of trainees infectious upon arrival or portion of staff infected off base on outbreak size before or after July 1, 2021**. (A) Portion of all individuals infected during training camp starting either before or after 07/01/21 as a function of the number of trainees who were infected in the two weeks prior to arrival. (B) Portion of all individuals infected during training camp starting either before or after 07/01/21 as a function of the number of staff who were infected off base.

