## Supplemental Information for "Community incidence patterns drive the risk of SARS-CoV-2 outbreaks and alter intervention impacts in a high-risk institutional setting"

**Table S1. Model parameters.**

| Parameter | Value | Reference |
|---|---|---|
| Incubation period (shape) | 5.807 | (*45*) |
| Incubation period (scale) | 0.948 | (*45*) |
| Duration of symptoms (d) | 10 | (*49*) |
| Proportion symptomatic (unvaccinated) | 0.31 | see note 1 |
| Proportion symptomatic (vaccinated) | 0.17 | see note 2 |
| Generation interval (shape) | 2.89 | (*32*) |
| Generation interval (scale) | 5.67 | (*32*) |
| Test specificity (PCR) | 0.998 | (*58*) |
| Test sensitivity (PCR) | 0.859 | (*58*) |

| | | |
|---|---|---|
| Test specificity (antigen) | 0.998 | (*58*) |
| Test sensitivity (antigen) | 0.859 | (*58*) |
| Protection from face masks (odds ratio) | 0.3 | (*59*) |
| Isolation length | 10 d | (*60*) |
| Relative infectiousness of asymptomatics | 0.35 | (*21*) |
| Immunity decay start (d) | 90 | (*50*) |
| Immunity decay rate ($d^{-1}$) | 0.0017 | (*50*) |
| Vaccine protection from infection (1st dose) | 0.368 | (*51*) |
| Vaccine protection from infection (2nd dose) | 0.775 | (*51*) |
| Vaccine protection from infection (booster dose) | 0.775 | *assumed* |

1. Based on an unpublished study of new military recruits during training camp in 2021. 2. Probability of a symptomatic infection for vaccinated individuals assuming that they are infected (protection against disease/protection against infection), adjusted for the ratio of symptomatic infection among 20-29 year olds relative to the overall population.

**Table S2. Variant-specific model parameters.** Relative transmissibility is transmissibility of a variant relative to the original SARS-CoV-2 strain. Relative cross-protection is the amount of cross-protection against a particular variant that is provided by a past infection with a different variant, relative to the level of protection provided by a same-variant infection. Vaccine protection is the level of protection against infection by a particular variant relative to vaccine-derived protection against infection by the original SARS-CoV-2 strain.

| Variant | Relative transmissibility | Source | Relative cross-protection | Source | Vaccine protection | Source |
|---|---|---|---|---|---|---|

| | | | | | | |
|---|---|---|---|---|---|---|
| Alpha | 1.29 | (*61*) | 0.95 | *assumed* | 0.95 | *assumed* |
| Gamma | 1.38 | (*61*) | 0.9 | *assumed* | 0.9 | *assumed* |
| Delta | 1.97 | (*61*) | 0.9 | *assumed* | 0.9 | *assumed* |
| Omicron | 2.85 | (*52*),(*53*) | 0.56 | (*53*) | 0.82 | (*62*) |

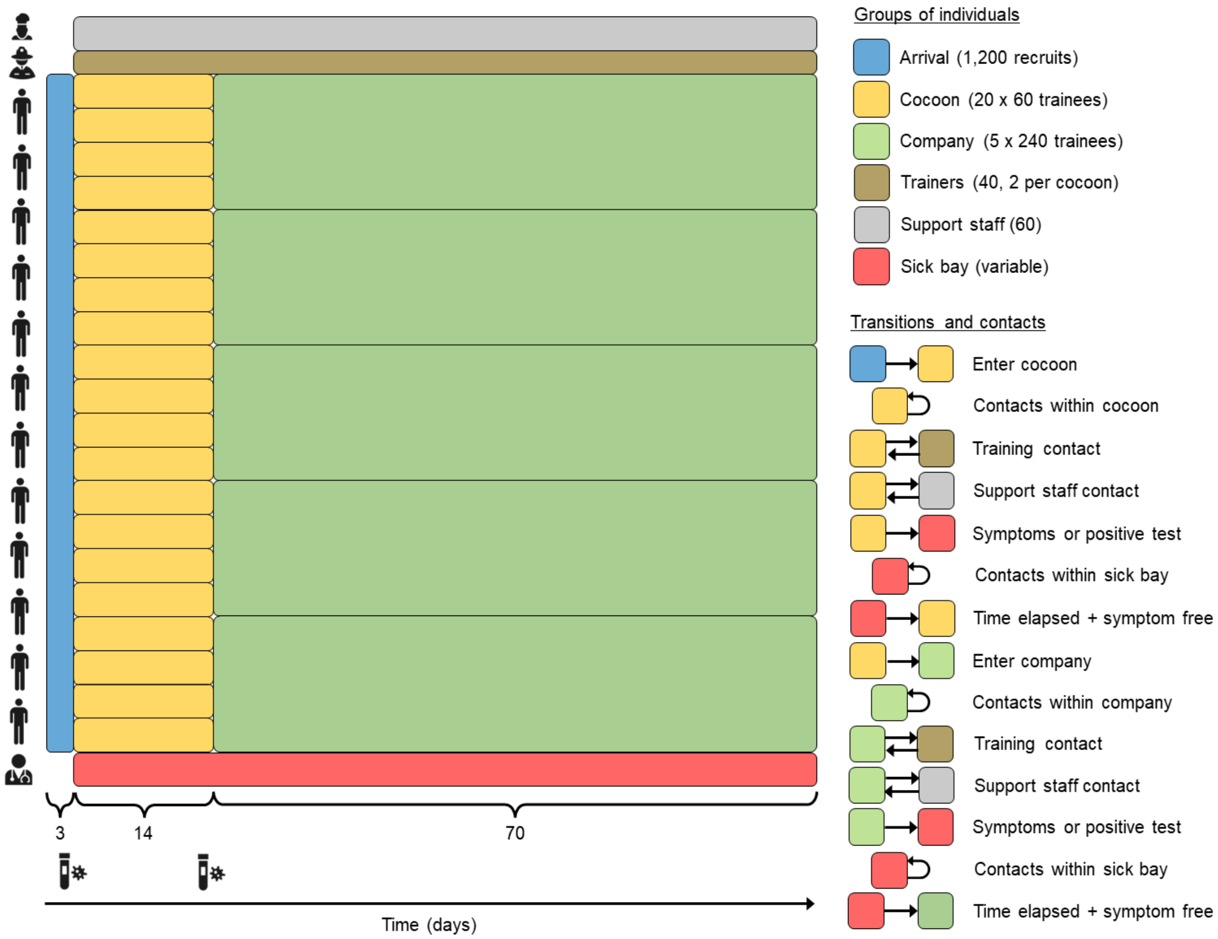


**Figure S1. Model schematic.** Trainees arrive at the beginning of training camp (blue), progress to cocoons of 60 trainees each for 14 days (yellow), and then progress to companies of 240 trainees each for 56 days (green). Trainees have contact with other trainees in their cocoon or company, with trainers (brown) assigned to their unit (two per cocoon, eight per company), and with support staff (gray). Trainees who test positive following arrival testing or presentation with symptoms are placed in the sick bay (red) for ten days (or until symptoms resolve) before returning to their unit. Trainers and support staff who test positive following presentation with symptoms isolate from home for a minimum of ten days. All processes in the model are defined on a daily time step.

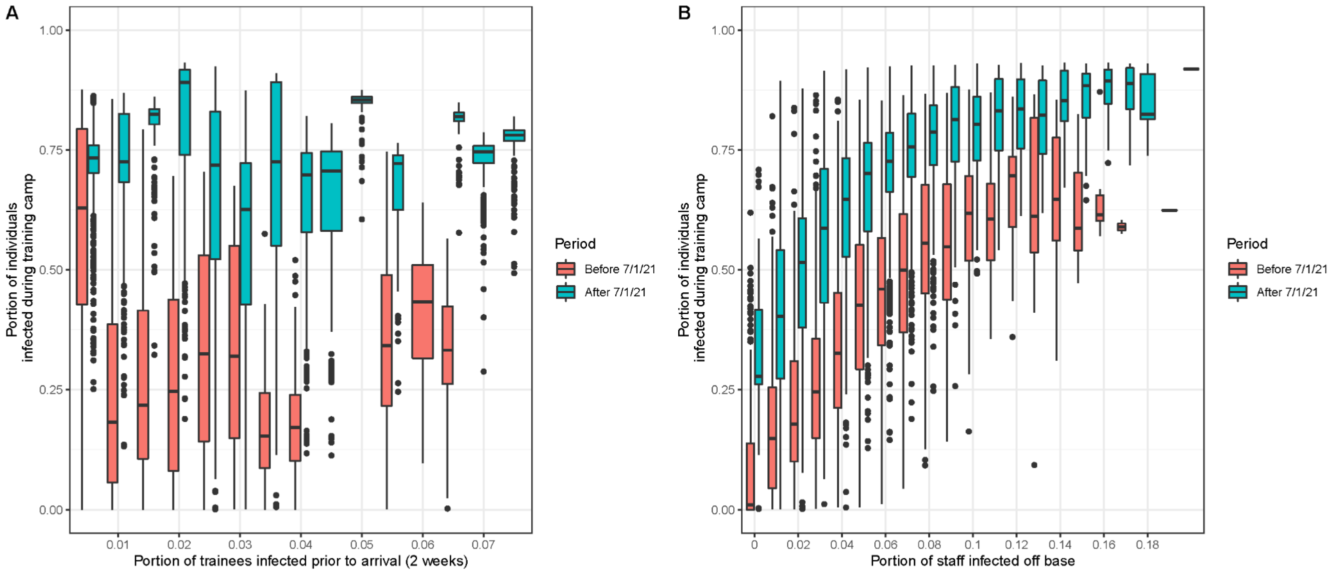


**Figure S2. Effect of portion of trainees infectious upon arrival or portion of staff infected off base on outbreak size before or after July 1, 2021.** (A) Portion of all individuals infected during training camp starting either before or after 07/01/21 as a function of the number of trainees who were infected in the two weeks prior to arrival. (B) Portion of all individuals infected during training camp starting either before or after 07/01/21 as a function of the number of staff who were infected off base.
